# Fasting C-peptide in Gestational Diabetes Mellitus

**DOI:** 10.1101/2024.08.27.24312520

**Authors:** Md. Rakibul Hasan, Nusrat Sultana, Mashfiqul Hasan, Sharmin Jahan, Muhammad Abul Hasanat

## Abstract

**Introduction:** Insulin secretion and sensitivity are thought to vary in gestational diabetes mellitus (GDM) as well as in different trimesters within the same individual.

**Objectives:** To see the fasting serum C-peptide in subjects with GDM as an indicator of basal insulin secretion and to compare it to the normal glucose tolerance (NGT) control.

**Method:** This case-control study investigated and compared fasting serum C-peptide in 35 GDM subjects with 37, age, BMI, and trimester-matched control. GDM was diagnosed by WHO 2013 criteria. Plasma glucose was measured by the glucose oxidase method and C-peptide by a chemiluminescent immunometric assay.

**Result:** Fasting serum C-peptides were significantly higher in the GDM group than that of the NGT (NGT vs. GDM: 1.45±0.76 vs. 1.99±1.28, mean±SD, p=0.032), but no difference was observed between GDM and NGT with a BMI <25kg/m^2^ (NGT vs. GDM: 1.25±0.44 vs. 1.37±0.77, mean±SD, p=0.597). Within the NGT group, fasting C-peptide was similar with a BMI cut-off 25kg/m^2^ (<25 vs. ≥ 25: 1.25±0.44 vs. 1.61±0.94, p=0.159); however, within the GDM group, the fasting C-peptide was significantly higher with a BMI ≥25 kg/m^2^ (<25 vs. ≥ 25: 1.37±0.77 vs. 2.45±1.41, mean ±SD, p=0.011). We did not observe any trimester-specific difference in fasting C-peptide between the NGT and the GDM (NGT vs. GDM: 1.55±0.45 vs. 1.67±0.57, p=0.736; 1.43±0.95 vs. 2.13±1.10, p=0.085; 1.43±0.71 vs. 1.97±1.60, p= 0.204; ng/dl, mean ± SD, for 1^st^, 2^nd^, and 3^rd^ trimester, respectively). Pearson correlation revealed BMI (r=0.389, p=0.001), Systolic BP (r=0.531, p<0.001), Diastolic BP (r=0.355, p=0.002), FPG (r=0.32, p=0.006) and 1hr PG on OGTT (r=0.423, p<0.001) had significant positive correlation with fasting C-peptide.

**Conclusion:** Fasting C-peptide increased significantly in GDM with BMI ≥25kg/m^2^, but not with <25kg/m^2^

## Introduction

Hyperglycemia in pregnancy is a very commonly encountered endocrine disorder in antenatal clinics all over the world, affecting about one in six pregnancies globally and one in four pregnancies in Southeast Asia. More than 80% of hyperglycemia in pregnancy is caused by gestational diabetes mellitus.^1^ Globally; its prevalence has been increasing over the last few decades. Pregnancy is a state of insulin resistance, to maintain blood glucose within normal range, beta cell of the pancreas increases its insulin secretion. GDM starts to develop when secretory reserve of beta cell can no longer compensate for increased insulin demand conferred by heightened insulin resistance. Although, insulin resistance is the most crucial in development of GDM, but impaired β-cell function and insulin secretory defects are also responsible for GDM in lean mothers.^2-3^ Impaired insulin secretion accounts for as much as 40% of gestational diabetes mellitus in leaner women in Japan.^4^ Maturity-onset diabetes of the young (MODY) is one of the leading causes of low insulin secretion. MODY gene mutation has been found in a significant proportion of pregnant women with GDM and young-onset diabetes patients. ^5-6^ Pregnant women with GDM who carried the risk allele (T) in TCF7L2 rs7903146 displayed significantly lower fasting insulin levels than those in non-carriers. ^7^ Early marriage and conception at a young age are very common in many developing countries. In our population, the majority of pregnant women are under the 30-year age group.^8^Therefore, there is a high possibility of the presence of a significant number of GDM mothers with low insulin secretion rather than insulin resistance as a major determinant. Screening of islets cell autoantibody, serum insulin, serum c-peptide, and genetic screening may help to identity the underlying causes of GDM but doing all these investigations are not feasible in a low-resource setting; furthermore, not all tests are available everywhere. Measuring fasting C-peptide may be a simple and good alternative for initial screening for insulin secretory status. Fasting serum C peptide is an important marker of basal insulin secretion. C peptide is secreted at equimolar concentration with insulin from beta cells of the pancreas. Insulin has a very short half-life compared to C peptide (3-5 minutes vs. 20–30 minutes), which affords a more stable test window for fluctuating beta cell responses. Half of all insulin secreted by the pancreatic beta cell is metabolized in the liver by first-pass metabolism; whereas C-peptide has negligible hepatic clearance. C peptide is cleared in the peripheral circulation at a constant rate, whereas insulin is cleared variably, making direct measurement of serum insulin less consistent. ^9^ This study aimed to see fasting serum C-peptide as an indicator of basal insulin secretion among GDM mothers and compare it with NGT control.

## Materials and Methods

### Study subjects and design

This case-control study was conducted in the Department of Endocrinology, Bangabandhu Sheikh Mujib Medical University (BSMMU), Dhaka. Pregnant women with ages ≥18 to 40 years, gestational weeks 6 to 38, were recruited from the antenatal clinic, Department of Gynecology & Obstetrics, BSMMU, from March 2016 to February 2017 after approval by the Institutional Review Board (IRB). Blood samples for OGTT and fasting serum C-peptide were collected at the GDM clinic of the endocrinology department of BSMMU. Blood was centrifuged immediately after collection at the GDM clinic and transported to the biochemistry and immunology laboratory within 30 minutes of collection for measurement of plasma glucose and serum C-peptide. A total of 35 GDM mothers with a 37,and 37 age, BMI, and trimester-matched pregnant women with NGT were included in this study. Pregnancy-specific WHO 2013 criteria were used to define them as either GDM or NGT. Demographic and clinical variables, including height, weight, body mass index (BMI, kg/m^2^), and blood pressure (mmHg), were recorded in a structured data collection sheet for analysis. Height was measured by using a stadiometer while standing upright on a flat surface without shoes. Weight was measured with a balance on a hard, flat surface. Blood pressure (BP) was measured in millimeters of mercury by a standard sphygmomanometer.

### Analytic method

C-peptide was measured by a chemiluminescent immunometric assay (Siemens 2008, C-peptide, IMMULITE 2000). The reference range of serum fasting C-peptide was 0.9–7.1 ng/mL. Serum glucose was measured by the glucose oxidase method in an automated analyzer [RA-50 analyzer (Dade Behring, Germany)].

### Statistical analysis

Quantitative data were expressed as mean±SD, whereas qualitative data were expressed as frequency distribution and percentage. Statistical analyses were performed using IBM SPSS Statistics for Windows version 22.0 (IBM Corp., Armonk, NY, USA). The association between categorical variables was analyzed by the chi-square (χ2) test and the continuous variable by the Student’s t-test. The correlation was done by Pearson’s correlation test. For all statistical tests, a P-value of less than 0.05 was considered statistically significant.

## Result

The majority of our study population (60%) in both groups were multigravida, and most of them were housewives (NGT vs. GDM: 81.1% vs. 68.6%, X^2^=1.505, p = 0.471). The baseline characteristics of the study subjects were similar between the NGT and GDM groups (Table 1). In general, we observed fasting C-peptides were significantly higher in the GDM group than that of the NGT group, but it did not significantly differ from that of the NGT group using the BMI cut-off of 25 kg/m^2^ (Table 2). Within the NGT group, fasting C-peptide was also similar compared to the BMI cut-off of 25 (<25 vs. ≥25 kg/m^2^; 1.25±0.44 vs. 1.61±0.94, p=0.159). However, within the GDM group, fasting C-peptide was statistically significantly higher in the GDM group with increasing BMI (<25 vs. ≥ 25 kg/m2: 1.37±0.77 vs. 2.45±1.41, p=0.011). We observed that GDM mothers had similar basal insulin secretion to NGT mothers when compared with BMI cut-offs of <25 kg/m2, but family history of diabetes mellitus among first-degree relatives was higher among GDM mothers with the same BMI category (Table 2), which may be related to genetic factors that may cause poor insulin secretion in GDM mothers with a lower BMI. We did not observe any trimester-specific difference in fasting C-peptide between the NGT and GDM groups (Table 3). Pearson correlation revealed BMI, systolic BP, diastolic BP, FPG, and 1 hour PG on OGTT had a significant positive correlation with fasting C-peptide, but age, gravida, gestational weeks, and 2 hours PG on OGTT had no relation with fasting C-peptide secretion (Table 4).

**Table 1:**
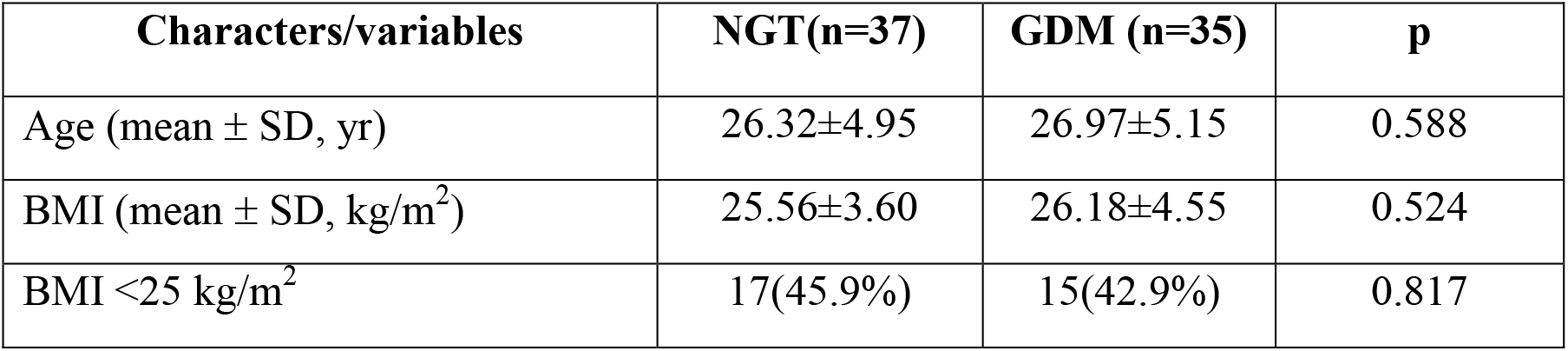
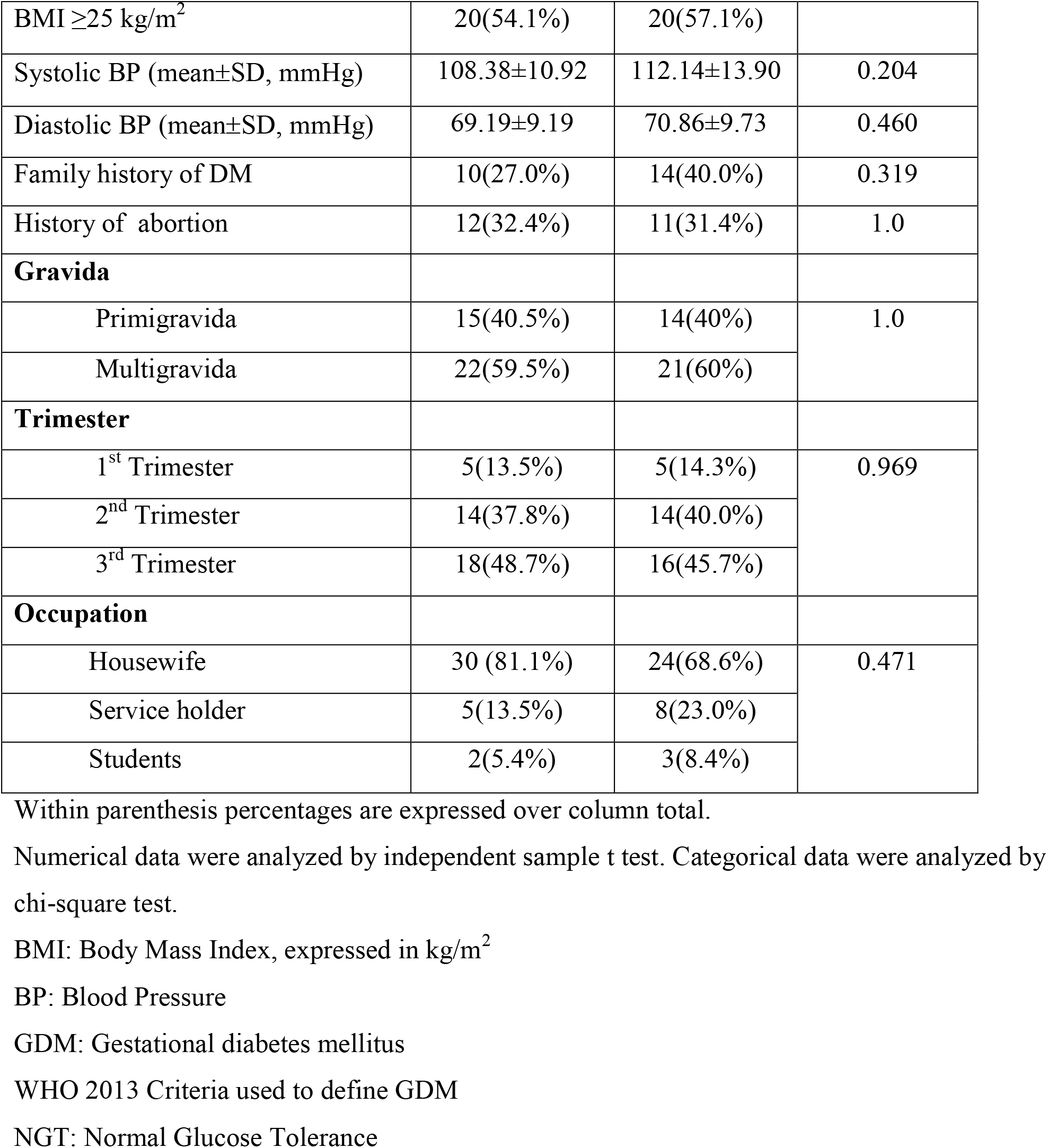
Characteristics of study subjects based on glycemic profile (N=72)

| Characters/variables | NGT(n=37) | GDM (n=35) | p |
| --- | --- | --- | --- |
| Age (mean ± SD, yr) | 26.32±4.95 | 26.97±5.15 | 0.588 |
| BMI (mean ± SD, kg/m<sup>2</sup>) | 25.56±3.60 | 26.18±4.55 | 0.524 |
| BMI <25 kg/m<sup>2</sup> | 17(45.9%) | 15(42.9%) | 0.817 |
| BMI $\geq 25$ kg/m<sup>2</sup> | 20(54.1%) | 20(57.1%) | |
| Systolic BP (mean±SD, mmHg) | 108.38±10.92 | 112.14±13.90 | 0.204 |
| Diastolic BP (mean±SD, mmHg) | 69.19±9.19 | 70.86±9.73 | 0.460 |
| Family history of DM | 10(27.0%) | 14(40.0%) | 0.319 |
| History of abortion | 12(32.4%) | 11(31.4%) | 1.0 |
| **Gravida** |  |  |  |
| Primigravida | 15(40.5%) | 14(40%) | 1.0 |
| Multigravida | 22(59.5%) | 21(60%) |  |
| **Trimester** |  |  |  |
| 1<sup>st</sup> Trimester | 5(13.5%) | 5(14.3%) | 0.969 |
| 2<sup>nd</sup> Trimester | 14(37.8%) | 14(40.0%) |  |
| 3<sup>rd</sup> Trimester | 18(48.7%) | 16(45.7%) |  |
| **Occupation** |  |  |  |
| Housewife | 30 (81.1%) | 24(68.6%) | 0.471 |
| Service holder | 5(13.5%) | 8(23.0%) |  |
| Students | 2(5.4%) | 3(8.4%) |  |

Within parenthesis percentages are expressed over column total.
Numerical data were analyzed by independent sample t test. Categorical data were analyzed by chi-square test.
BMI: Body Mass Index, expressed in kg/m<sup>2</sup>
BP: Blood Pressure
GDM: Gestational diabetes mellitus
WHO 2013 Criteria used to define GDM
NGT: Normal Glucose Tolerance

**Table 2:**
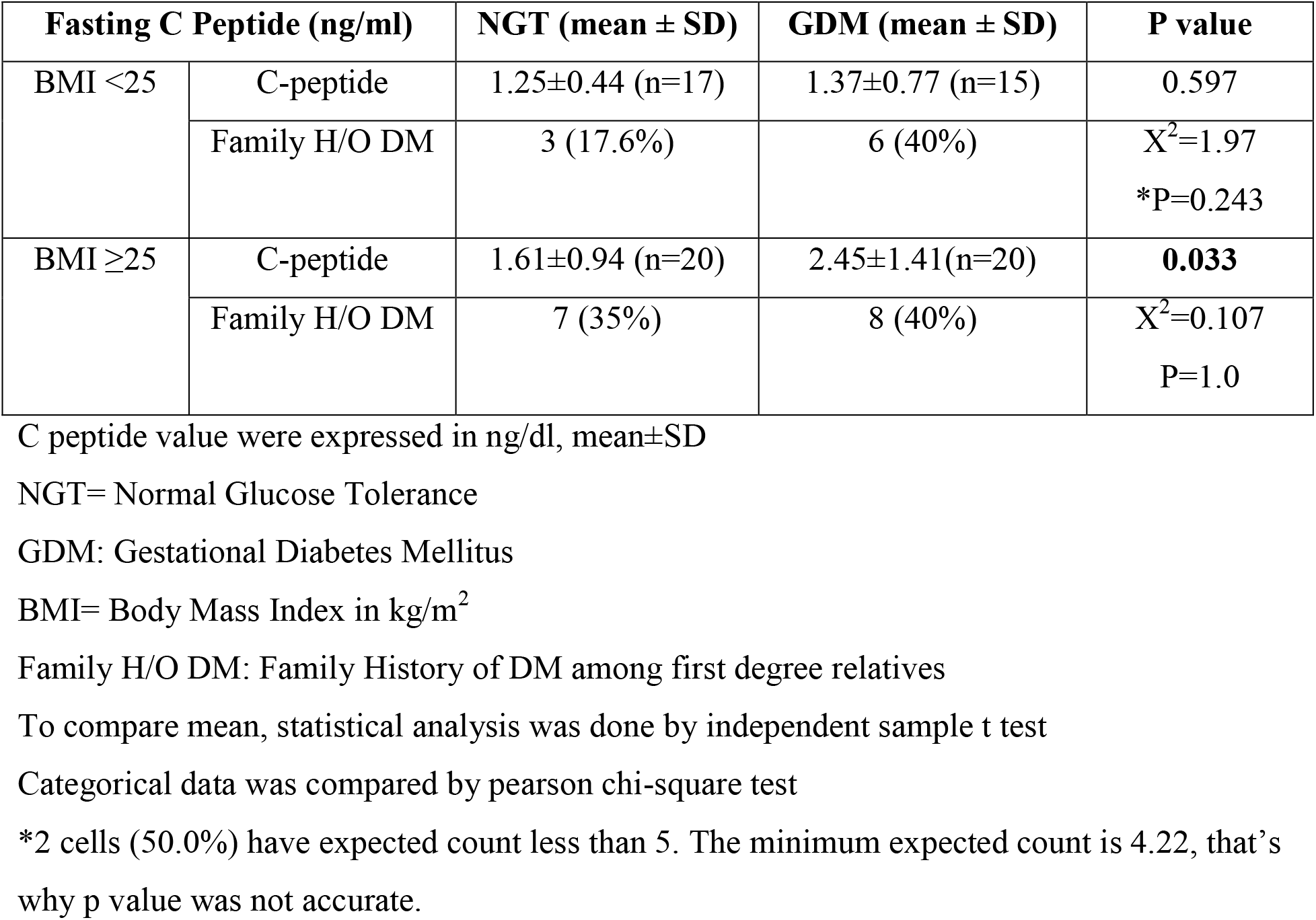
Comparison of Fasting C peptide between NGT vs GDM (N=72) at different BMI.

| Fasting C Peptide (ng/ml) | | NGT (mean ± SD) | GDM (mean ± SD) | P value |
| --- | --- | --- | --- | --- |
| BMI <25 | C-peptide | 1.25±0.44 (n=17) | 1.37±0.77 (n=15) | 0.597 |
|  | Family H/O DM | 3 (17.6%) | 6 (40%) | X<sup>2</sup>=1.97<br>*P=0.243 |
| BMI ≥25 | C-peptide | 1.61±0.94 (n=20) | 2.45±1.41(n=20) | **0.033** |
|  | Family H/O DM | 7 (35%) | 8 (40%) | X<sup>2</sup>=0.107<br>P=1.0 |

C peptide value were expressed in ng/dl, mean ± SD
NGT= Normal Glucose Tolerance
GDM: Gestational Diabetes Mellitus
BMI= Body Mass Index in kg/m<sup>2</sup>
Family H/O DM: Family History of DM among first degree relatives
To compare mean, statistical analysis was done by independent sample t test
Categorical data was compared by pearson chi-square test
\*2 cells (50.0%) have expected count less than 5. The minimum expected count is 4.22, that's why p value was not accurate.

**Table 3:**
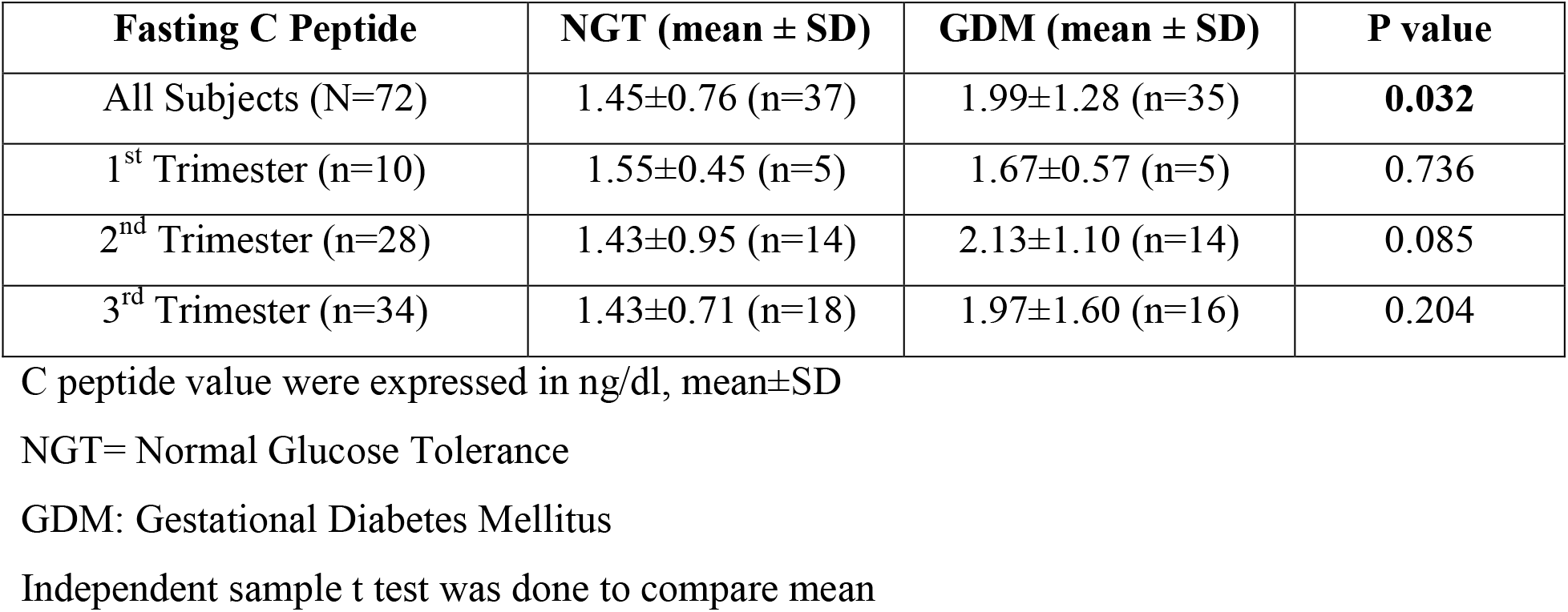
Comparison of Fasting C peptide between NGT vs. GDM (N=72) at different Trimesters.

**Table 4:** Pearson Correlation of Fasting C-Peptide with various factors of study subjects.

| Determinant | Correlation coefficient r | P value |
| --- | --- | --- |
| C-peptide vs. Age | 0.108 | 0.369 |
| C-peptide vs. BMI | **0.389** | **0.001** |
| C-peptide vs. SBP | **0.531** | **<0.001** |
| C-peptide vs. DBP | **0.355** | **0.002** |
| C-peptide vs. Week of gestation | -0.032 | 0.789 |
| C-peptide vs. Gravida | -0.081 | 0.501 |
| C-peptide vs. FPG | **0.321** | **0.006** |
| C-peptide vs. 1h PG | **0.423** | **<0.001** |
| C-peptide vs. 2h PG | 0.171 | 0.150 |

BMI= Body Mass Index in $\text{kg/m}^2$
SBP: Systolic Blood Pressure
DPB: Diastolic Blood Pressure
FPG: Fasting Plasma Glucose
1h PG: Plasma Glucose at 01hour of OGTT
2h PG: Plasma Glucose at 02hour of OGTT

## Discussion

The fasting C-peptide is an indicator of basal insulin secretion. In our study, we observed that fasting C-peptide was significantly higher in the GDM group than that in the NGT group. However, this difference was not observed when we compared C-peptide between GDM and NGT with a BMI <25kg/m^2^. Fasting C-peptide did not significantly differ in the NGT group based on BMI category (<25 vs. ≥25kg/m^2^); in the GDM group, fasting C-peptide increased significantly with increasing BMI (≥25kg/m^2^). These findings indicate that insulin secretion is increased in GDM with an increasing BMI. However, the subjects who were diagnosed with GDM with a relatively lower BMI had no significantly increased insulin secretion compared to the BMI-matched NGT, despite having higher plasma glucose. This group of GDM mothers might have insulin secretory defects that lead to the development of gestational diabetes mellitus.

Fakhrul-Alam M. et al. reported the same findings. He assessed the insulin sensitivity and secretory capacity of the GDM mother with age and a BMI-matched control. ^3^ He observed that insulin secretion was statistically significantly low in GDM mothers compared to NGT mothers with a BMI cut point of <23 kg/m^2^. Tania et al.also observed that the fasting C peptide in GDM mothers was also statistically significantly lower in lean GDM mothers compared to NGT. ^10^ In our study, it was not low; rather, it was statistically similar between GDM and NGT. This may be due to using a BMI cut-off of 25 kg/m^2^ in our study, whereas it was 23 kg/m^2^ in the previous study. In our study, we did not observe any significant difference in fasting C peptide in the NGT group based on BMI category (≥25 vs. <25 kg/m^2^). However, ElżbietaPoniedziałek-Czajkowska et al.observed significantly higher concentrations of C-peptide (2.77 ± 1.88 vs. 2.25 ± 1.42 ng/mL, p = 0.034) in the obese NGT group than that of the normal BMI NGT group. This may be due to the very high mean BMI in the obese NGT group compared to the normal BMI group (32.68 ± 5.12 vs. 21.07 ± 2.13, mean±SD, kg/m^2^), which was not very high in our study. ^11^ In another study from China, fasting C peptide was similar between the GDM and NGT groups during the last trimester of pregnancy and after delivery. We also observed no trimester-specific difference in fasting C peptide between the GDM and NGT mothers. However, the overall fasting C peptide in the GDM group was significantly higher than that of the NGT. There are many causes of insulin secretory defects. Maturity-onset diabetes in the young (MODY) gene mutation is one of the important regions of defective insulin secretion. In one study, screening of GCK, HNF1A, HNF4A, INS, and HNF1B gene mutations in 354 GDM women revealed a significant number of GDM results from these gene mutations.They reported a 5.9% prevalence of possible diabetes-predisposing gene variants. Overall, 11% of the total number of diabetic women at follow-up had diabetes-predisposing variants in GCK, HNF1A, HNF4A, HNF1B, or INS. ^13^ In another study, the prevalence of MODY gene mutation was 12.6% among South Asian individuals who were referred for MODY testing. ^14^ A genetic polymorphism of TCF7L2 is also an important cause of defective insulin secretion. The frequency of polymorphism of TCF7L2 was observed to be higher in lean and young-onset GDM mothers studied in Bangladeshi GDM women. The CT/TT genotype was associated with a higher frequency of GDM in women aged <25 years (CT/TT vs. CC: 58.3% vs. 17.4%, p = 0.022) and BMI <25 kg/m^2^ (CT/TT vs. CC: 61.5% vs. 18.2%, p = 0.024). ^15^ Our study assessed fasting C-peptide in GDM as an indicator of basal insulin secretion. We observed similar insulin secretion in GDM mothers compared with NGT mothers with a BMI cut of 25 kg/m^2^. Our study findings help us to rethink the pathogenesis of GDM in normal-weight pregnant women and guide us for further research on this topic.

## Data Availability

All data produced in the present study are available upon reasonable request to the authors

## Declarations

## Acknowledgments

We are grateful to the study participants and their attendants.

## Data Availability

Any inquiries regarding supporting data availability of this study should be directed to the corresponding author and are available from the corresponding author upon reasonable request.

## Ethics Approval and Consent to Participate

This study was approved by the Institutional Review Board (IRB) of BSMMU, Reg: No. BSMMU/2016/2746, approved on 08-03-2016. All procedures performed in this study involving human participants were following the ethical standards of the IRB and with the 1964 Helsinki Declaration and its later amendments or comparable ethical standards. Informed written consent was obtained from each of the participants included in the study.

